# Automating the generation of antimicrobial resistance surveillance reports: a proof-of-concept study in seven hospitals in seven countries

**DOI:** 10.1101/2020.04.27.20072025

**Authors:** Cherry Lim, Thyl Miliya, Vilada Chansamouth, Myint Thazin Aung, Abhilasha Karkey, Prapit Teparrukkul, Rahul Batra, Lan Nguyen Phu Huong, John Stelling, Paul Turner, Elizabeth Ashley, Rogier H van Doorn, Htet Naing Lin, Clare Ling, Soawapak Hinjoy, Sopon Iamsirithaworn, Susanna J Dunachie, Tri Wangrangsimakul, Viriya Hantrakun, William Schilling, Yen Lam Minh, Tan Le Van, Htay Htay Hlaing, Mayfong Mayxay, Manivanh Vongsouvath, Buddha Basnyat, Jonathan Edgeworth, Sharon J Peacock, Guy Thwaites, Nicholas PJ Day, Ben S Cooper, Direk Limmathurotsakul

**Author notes:** **Corresponding author:** Cherry Lim. Mahidol Oxford Tropical Medicine Research Unit, Faculty of Tropical Medicine, Mahidol University, 420/6 Rajvithi Road, Bangkok, 10400, Thailand. **Alternative Corresponding author:** Direk Limmathurotsakul. Mahidol Oxford Tropical Medicine Research Unit, Faculty of Tropical Medicine, Mahidol University, 420/6 Rajvithi Road, Bangkok, 10400, Thailand.

## Abstract

**Background:** Reporting cumulative antimicrobial susceptibility testing data on a regular basis is crucial to inform antimicrobial resistance (AMR) action plans at local, national and global levels. However, analysing data and generating a report are time-consuming and often require trained personnel. We illustrate the development and utility of an offline, open-access and automated tool that can support the generation of AMR surveillance reports promptly at the local level.

**Methods:** An offline application to generate standardized AMR surveillance reports from routinely available microbiology and hospital data files was written in the R programming language. The application can be run by a double-click on the application file without any further user input. The data analysis procedure and report content were developed based on the recommendations of the World Health Organization Global Antimicrobial Resistance Surveillance System (WHO GLASS). The application was tested in Microsoft Windows 10 and 7 using open-access example data sets. We then independently tested the application in seven hospitals in Cambodia, Lao People’s Democratic Republic (PDR), Myanmar, Nepal, Thailand, the United Kingdom, and Vietnam.

**Findings:** We developed the AutoMated tool for Antimicrobial resistance Surveillance System (AMASS), which can support clinical microbiology laboratories to analyse their microbiology and hospital data files (in CSV or Excel format) onsite and promptly generate AMR surveillance reports (in PDF and Excel formats). The data files could be those exported from WHONET and/or other laboratory information systems. The automatically generated reports contain only summary data without patient identifiers. The AMASS application is downloadable from www.amass.website. The participating hospitals tested the application and deposited their AMR surveillance reports in an open-access data repository.

**Interpretation:** The AMASS application can be a useful tool to support the generation and sharing of AMR surveillance reports.

**Funding:** Mahidol Oxford Tropical Medicine Research Unit (MORU) is funded by the Wellcome Trust (Grant no. 106698/Z/14/Z). Oxford University Clinical Research Unit (OUCRU) is funded by the Wellcome Trust (Grant no. 106680/B/14/Z). The investigators are funded by the Wellcome Trust (CL is funded by a Training Research Fellowship [Grant no. 206736] and DL is funded by an Intermediate Training Fellowship [Grant no. 101103]). BSC is funded by the UK Medical Research Council and Department for International Development (Grant no. MR/K006924/1). The funder has no role in the design and conduct of the study, data collection, or analysis and interpretation of the data.

## INTRODUCTION

Generating and sharing antimicrobial resistance (AMR) surveillance reports are fundamental elements of actions against AMR infections at local, national and international levels. Information on patterns of antimicrobial susceptibility is important to guide empiric choice of therapy, monitor resistance trends and detect outbreaks of AMR infections at the local level [1–3]. Combining data and reports at the national level provides evidence to inform the implementation of national action plans, decide on resource allocation for interventions, and monitor the impact of those interventions [3–5]. The Review on AMR chaired by Jim O’Neill estimated that 700,000 global deaths are attributable to AMR infections each year (including bacterial infections, HIV, TB and malaria) [6], and had an enormous global impact [7,8]. While this represents a very rough estimate subject to well-documented limitations [1,9,10], importantly the report highlighted the need for improved AMR surveillance.

Methods to analyse data and generate AMR surveillance reports are gradually being standardised worldwide [3,11–15]. Recently, the World Health Organization (WHO) launched the Global Antimicrobial Resistance Surveillance System (GLASS) attempting to standardize the AMR surveillance data reporting format [11,16,17]. In general, it is recommended that (a) duplicated results on bacterial isolates should be removed, and (b) data should be stratified by origin of infection (community or hospital) whenever possible [11,15]. A simple de-duplication process is to include only the first isolate of a species per patient per specimen type per survey period in the report [3,11]. The origin of infection is defined by using specimen collection date and hospital admission date as a proxy to define where the infection was likely contracted (community or hospital) [3,11].

Even in areas where AMR surveillance data are available, there are many barriers to utilizing such data [18]. Many hospitals in low and middle-income countries (LMICs) lack the time and resources needed to analyse the data (particularly to de-duplicate and validate the accuracy of the summary data), write the reports, and to release the data or reports [18]. There are open-access Laboratory Information Systems (LIS) [19], and microbiology laboratory database softwares, including WHONET [20], that are useful for recording and analysing the data, and generating figures to support the generation of AMR surveillance reports for hospitals in LMICs. There are no freely-available automated system that can generate summary reports for immediate use and be operated by non-technical staff. In addition, to generate AMR surveillance reports stratified by the origin of infection, additional data on hospital admission are frequently needed [21–24]. This is because hospital admission dates are not generally collected in microbiology laboratory data. In many hospital settings microbiology and hospital admission data are held in separate computers or systems with restricted access. Even in high-income countries, many hospitals lack well-trained clinical microbiologists, epidemiologists or data experts with adequate skills in statistical software (such as R, SAS, SPSS and STATA) to merge and de-duplicate data in the separated databases and generate reports stratified by the origin of infection.

Here, we developed an application, termed AutoMated tool for Antimicrobial resistance Surveillance System (AMASS), which can support a local hospital to independently analyse routinely collected electronic data and rapidly generate AMR surveillance reports. We tested AMASS in seven hospitals in Cambodia, Lao PDR, Myanmar, Nepal, Thailand, the United Kingdom and Vietnam, and deposited the report from each hospital in an open-access platform.

## METHODS

### Design of the application

The tool operates by reading and processing the raw data files to automatically produce AMR surveillance reports. To ensure the tool is fully open access, we built the application in R (version 3.6.2), which is a free software environment. We then placed the application within a user-friendly interface, which only requires a double-click on the application file to run the automation without the need to understand the R program. We included both the R portable (version 3.4.3) and RStudio (version 1.1.423) within the downloadable package so that the application can run without the need to install R or any program prior to running the application.

### Data processing and statistical analysis

A schematic overview of input data, data processing, statistical analyses and output is shown in Supplementary Figure 1. In brief, input data are microbiology data files (in CSV or Excel formats) with or without hospital data files, which can be exported from WHONET or a LIS with data export capacity. The data processing and analysis algorithms, including data-deduplication, were developed based on the WHO GLASS recommendations [11,17]. The reports were designed to have a similar format to the WHO GLASS 2018 report [14].

The application de-duplicates the data by including only the first isolate per sample type per pathogen per survey period for each patient [11]. The application currently includes only blood specimens, and eight pathogens: namely *Acinetobacter* spp., *Escherichia coli*, *Enterococcus* spp., *Klebsiella pneumoniae*, *Salmonella* spp., *Staphylococcus aureus*, *Streptococcus pneumoniae*, and *Pseudomonas aeruginosa*. Both *Enterococcus* spp., and *P. aeruginosa* were added on top of the six priority pathogens described by the WHO GLASS for bacteraemia [17,25] because both pathogens are common causes of bacteraemia [15,21], and were included in the 2015 global priority list of AMR bacteria [25]. The list of pathogen-antibiotic combinations was modified from the WHO GLASS (Supplementary Table 1). Infections are stratified into community-origin or hospital-origin using hospital admission dates and specimen collection dates, when available [11,17]. Patients with the first specimen culture positive for the pathogen taken in the outpatient setting or on the first or second day of hospitalization are classified as having community-origin infection [11,17]. Patients with the first specimen culture positive for the pathogen taken on hospital day three or later are classified as having hospital-origin infection [11,17]. Alternatively, in cases where users have data on origin of infection assigned by the attending physicians or infection control team of the hospital, AMASS can instead use those categorizations to stratify the infection into community-origin or hospital-origin. Prevalence and incidence rates are estimated based on the recommendations of the WHO GLASS (Supplementary Text 1) [11,17]. An additional report on mortality involving AMR and non-AMR infections is generated when mortality data are available in the hospital admission data file. The term “mortality involving AMR and antimicrobial-susceptible infections” is used because the mortality reported in the hospital admission data are all-cause mortality [1]. This measure of mortality includes deaths caused or contributed by other underlying and intermediate causes. Therefore, the term “involving AMR infections” is used, in accordance with the term used by the UK Office for National Statistics [26,27].

**Table 1.** Features of the AMASS application.

| Features | Descriptions |
| --- | --- |
| **Open-access** | **AMASS is open-access and can be downloaded** from www.amass.website or https://figshare.com/articles/AMASS_zip/7763330 (zip file size 177.54MB) |
|  | **AMASS was developed by R, which is a free software.** In the download package for AMASS (AMASS.zip), there is a folder that contains R-Portable and RStudio, which supports the data processing and analysis to generate AMR surveillance report automatically. |
|  | **AMASS is under CC-BY 4.0 license.** Users can share (copy and redistribute the material in any medium or format) and modify the R codes of the AMASS under the terms & conditions of the Creative Commons license. |
| **User-friendly** | **AMASS can be run by a double-click on the application icon.** Data analysis and AMR surveillance report generation are automated by the AMASS application. |
|  | **Data cleaning, de-duplication, and analysis are performed rapidly** (it took about 1-3 minutes to automatically produce an AMR surveillance report using example data sets provided in the AMASS package). |
|  | **No additional program or software is needed.** All the essential software is stored in the AMASS package and will operate automatically after a double click the application file (AMASS.bat). |
|  | **Users do not need to understand R program or write any codes to run the AMASS application.** |
| **Highly-compatible** | **AMASS works with raw data files in either CSV or Excel format,** which can be commonly exported from WHONET and other software, program or data management systems used for microbiology data and hospital admission data. |
|  | **AMASS uses data dictionary files (in Excel format) to accommodate data exported from different software, program or systems** that may have different ways to name data variables and data values (Figure 2). |
|  | **AMASS dictionary files can be re-used by users in the future** (e.g. monthly, quarterly and yearly) if the structures of the new raw microbiology data file and hospital admission data file remain unchanged. |
|  | **AMASS uses a tier-based approach based on availability of raw data files to generate reports.** Users with limited data availabilities (e.g. microbiology data with only culture positive results) can still utilize de-duplication and report generation functions of the AMASS. Users with additional data (e.g. microbiology data with culture negative results and hospital admission data) will receive additional reports (e.g. sample-based surveillance report with stratification by infection origin). |
| **High data security** | **AMASS does not require the Internet to operate.** Users do not have to transfer raw individual data (which may contain identifiable information) to any institution outside |
|  | of the hospital to analyse the data and generate the reports. AMASS can be run within a standalone computer within the local hospital under the local data security. Hence, the AMASS does not increase any risks of breaching individual patient data confidentiality. |
| **Easy-to-use outputs** | **The automatically generated AMR surveillance report is in PDF format**, which is easy to print, read and share within and outside the hospitals. |
| **Easy-to-share outputs** | **The report (in PDF format) and aggregated summary data files (in CSV format) contains no individual level patient data**, and could be readily shared with national and international organizations. |

### Example data sets

Two example data sets are provided in the downloadable package of the AMASS application. The first example data set is the open-access demonstration file from WHONET [19]. The second example data set is a hypothetical data set generated for AMASS. The second example data set was created to represent a large data set from a 1000-bed hospital, containing both microbiology and hospital admission data. A detailed description on how the second example data set was hypothetically generated is in Supplementary Text 2. In brief, two synthetic data files (microbiology_data.xls and hospital_admission_data.xls files) were generated based on the summary data in the AMR surveillance report from 2015 of Sunpasitthiprasong Hospital, Ubon Ratchathani, Thailand. Variables in the microbiology data file include hospital number (HN), specimen type, specimen collection date, culture result, and antibiotic susceptibility testing result, and each row contains information for each specimen. Variables in the hospital admission data file include HN, age, sex, admission date, discharge date and in-hospital discharge outcome. All data were randomly generated using STATA 15.1 (Texas, USA).

### Testing AMASS using hospital data

Seven hospitals participated in the study: Angkor Hospital for Children in Cambodia, Mahosot Hospital in Lao PDR, North Okkalapa General and Teaching Hospital in Myanmar, Patan Hospital in Nepal, Sunpasitthiprasong Hospital in Thailand, St Thomas’ Hospital in the United Kingdom, and the Hospital for Tropical Diseases in Vietnam. The hospitals were selected because microbiology data are collected routinely, and they have prior experience in data quality controls. Moreover, the hospitals varied in LIS used for data storage and are good examples to demonstrate the practicality and usability of AMASS in different settings, to a wider audience. The study was approved by the Oxford Tropical Research Ethics Committee, University of Oxford, and local Ethics Committees. C.L. corresponded with the participating hospitals and demonstrated how the application operates using the example data files. The local hospital staff operated the application using their local data by themselves. All microbiology and hospital data were stored independently within their hospital computers under their local data protection standards. Automatically generated reports and anonymous summary data from each hospital are publicly available and deposited in data repositories with permission from each hospital.

## RESULTS

### Overview of AMASS

The AMASS application has been developed to support clinical microbiology laboratories to automatically analyse hospital local data files (in CSV or Excel formats) and generate AMR surveillance reports (in PDF and Excel formats) promptly. Six steps are followed to generate the AMR surveillance reports (Figure 1 and Supplementary video file 1). (1) Download the AMASS package from www.amass.website. (2) Obtain routinely collected raw microbiology data file and, if available, hospital admission data file, and then, save the data files in the folder of the AMASS application. (3) Configure data dictionaries. Two data dictionaries are provided to accommodate different ways of naming variables (e.g. sex or gender) and data values (e.g. M and F, or Male and Female). How the data dictionaries functioned is described in more detail in the “Input requirements” section. (4) Double-click on the AMASS.bat file to run the application. (5) Review and validate the AMR surveillance reports (generated in PDF format) and the anonymous summary data (generated in CSV format). (6) Share the reports within the hospital, especially with the local infection control team. The reports and anonymous summary data contain no patient identifiers. Therefore, users may share the reports and anonymous summary data with national and international organizations or make the reports and anonymous summary data open access.

**Figure 1.**
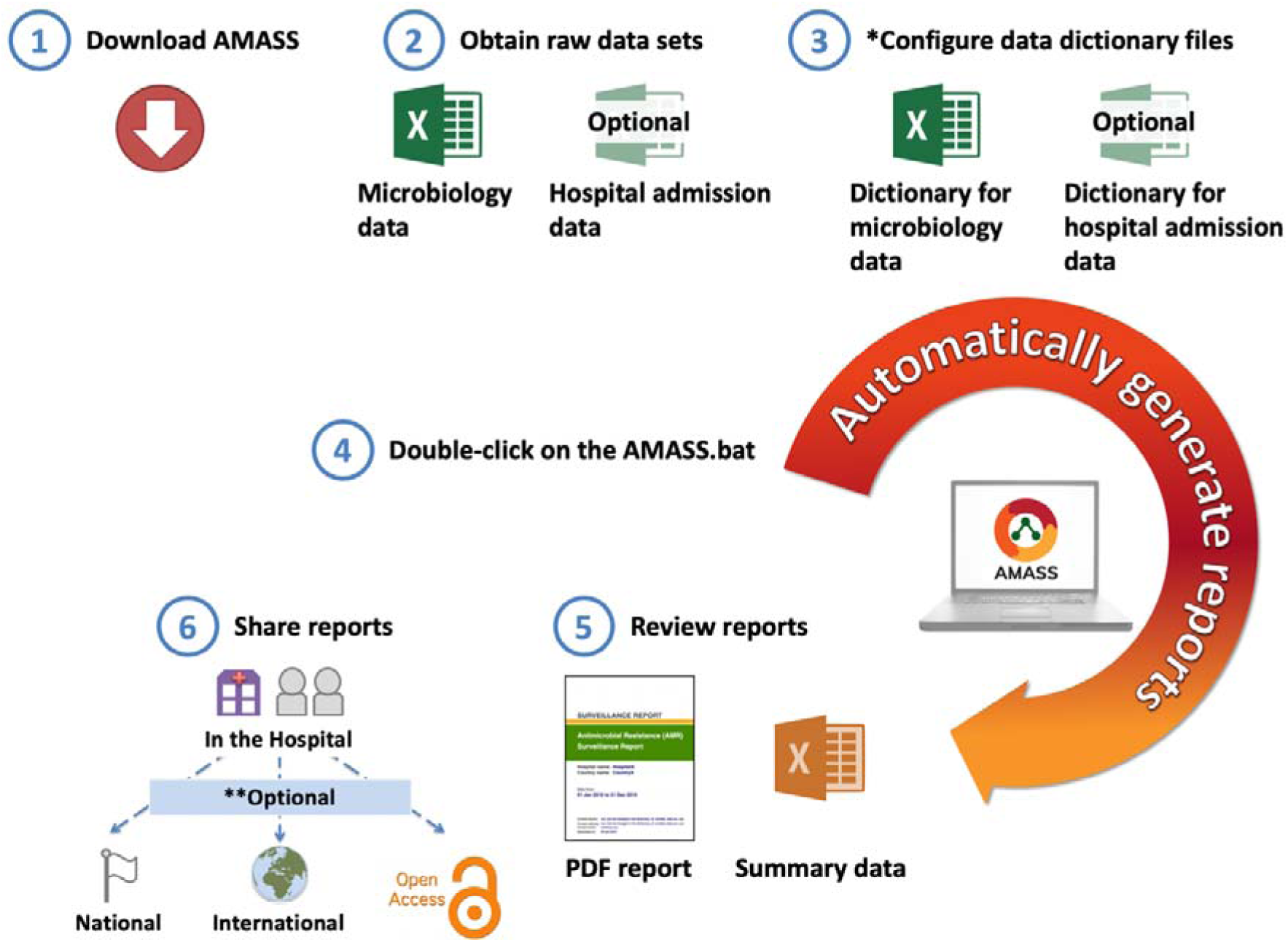
A conceptual flow of AutoMated tool for Antimicrobial resistance Surveillance System (AMASS). Footnote of Figure 1. Step 1 (Download AMASS) and step 3 (Configure data dictionary files) are one-time steps. Step 2 (obtain data), step 4 (run AMASS), step 5 (review report), and step 6 (share report) are ongoing steps that users could repeat regularly (i.e. monthly, quarterly). *Two data dictionary files (in Excel format) are provided to allow the application to understand how variables and values of each variable are named in the raw data files in different settings. Those data dictionary files can be re-used in the subsequent runs of AMASS, as long as how variables and values of each variable are named in the raw data files remains the same. Details on how to configure the data dictionary can be found in Figure 2 and Supplementary video 2. **The AMR surveillance report and summary data generated contain no patient identifiable information. The decision to share the report and summary data to national or international AMR organizations is solely up to the jurisdiction of the hospital.

### Features of AMASS

Key features of the AMASS are listed in Table 1. In brief, the AMASS 1) can be downloaded from www.amass.website and be used under Creative Commons Attribution 4.0 International license (i.e. it is open-access); 2) only requires users to double-click on the application icon to perform data processing and analysis (i.e. user-friendly); 3) reads CSV and EXCEL files that can be exported from WHONET, LIS, or electronic health record (eHR) systems (i.e. highly compatible); and 4) can run on stand-alone and offline computers in the hospitals under their local data protection standards. All data processing and report generation are done locally, no raw data are shared, and final reports and output summary data files contain no patient identifier (i.e. high data security).

Moreover, AMASS supports the rapid use of AMR surveillance data at the local level. The readily printable report in PDF format can be reviewed and validated by non-statisticians. When errors are found in the raw data files (e.g. incomplete data) or the data dictionary files (e.g. typos), the application can be promptly re-run after fixing the errors. The summary statistics shown in the PDF report are also saved in the aggregate summary data file (in common-separated value format) ready for re-use.

### Input requirements

To align with formats of commonly exported microbiology data and hospital admission data, AMASS reads data files in both CSV and Excel formats. The microbiology data needs to be in wide format, meaning that each row contains data from a single clinical sample. The key variables required for the microbiology data file include patient identifier, specimen collection date, specimen type, culture result, and antimicrobial susceptibility test interpretation per each antibiotic. The hospital admission data file is optional but, if available, also needs to be in wide format. The key variables needed for the hospital admission data file include patient identifier, admission date, gender, age, and, if possible, in-hospital discharge outcome. The current version of AMASS uses Gregorian calendar and required the date to be in the order of date, month, and year in any format (i.e. either text [English] or numeric).

There are two data dictionary files provided for the users to accommodate different ways of naming data variables and data values (Figure 2). The first data dictionary file (dictionary_for_microbiology_data.xls) is for the microbiology data file. For example, the AMASS uses the variable name “hospital_number” as a patient identifier (Row 3, Column A of the data dictionary file). In cases where a raw microbiology data file uses a different name for the patient identifier (e.g. hn), users would need to fill “hn” in the data dictionary file (Row 3, Column B of the data dictionary file). This allows the AMASS application to know that the variable “hn” of the raw microbiology data file is the patient identifier (i.e. “hospital_number”). The second data dictionary file (dictionary_for_hospital_admission_data.xls) is for the hospital admission data file, which is to be used likewise. Supplementary video 2 is a step-by-step tutorial on how to use and configure the data dictionaries.

**Figure 2.**
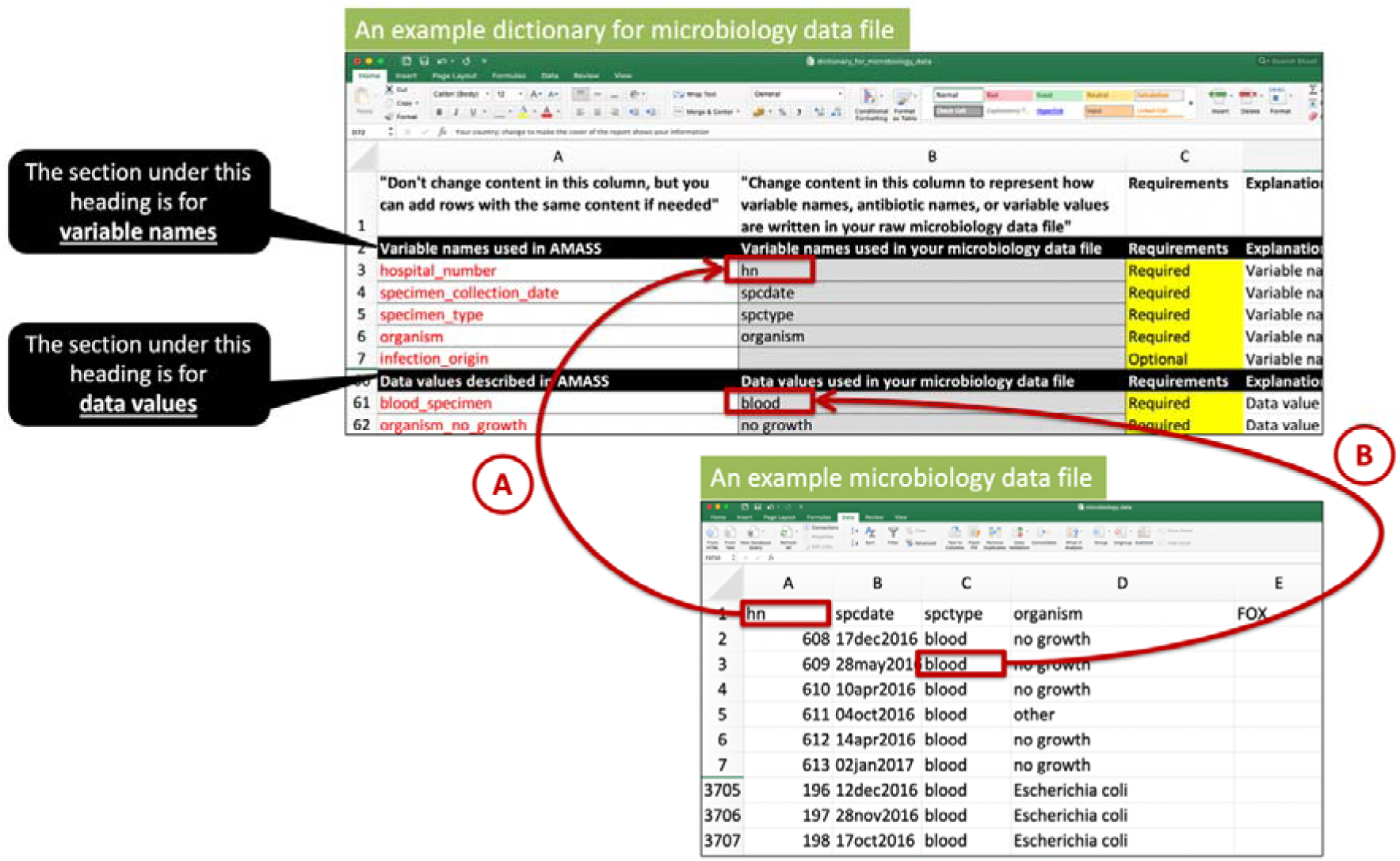
An example of how to complete a data dictionary file. Footnote of Figure 2. For a first-time user, the user may need to complete a data dictionary file, by filling in variables names used in their raw data files into the data dictionary files (e.g. arrow A). This is to allow AMASS to understand that the variable “hospital_number” used by AMASS is named as “hn” in the user’s raw microbiology data file. Then, users need to enter how data values are named in their raw data files (e.g. arrow B). This is to allow AMASS to understand that the data value named “blood_specimen” is named as “blood” in user’s raw microbiology data file. Please note that the contents in the first column of the data dictionary file must remain unchanged. Users can add new rows but the content in the cell in the first column must not be changed. For example, users can define that both “*E. coli* (ESBLs producing strain)” and “*Escherichia coli*” in their raw microbiology data file means “organism_escherichia_coli” by AMASS. The example data dictionary files shown in the figure are available in the Example_Dataset_2 folder (within the AMASS download package).

### Outputs generated by AMASS

We illustrate the AMR surveillance reports generated from AMASS using the two open-access example data sets provided within the download package. Supplementary file 3 and 4 contain AMR surveillance reports generated from the first and second example data sets, respectively. Supplementary video 1 illustrates how to test the AMASS using example data sets step-by-step.

In short, the automatically generated report on AMR surveillance contains six sections (Figure 3); including: [section one] data overview; [section two] an isolate-based report; [section three] an isolate-based report with stratification by origin of infection; [section four] a sample-based report; [section five] a sample-based report without stratification by infection; and [section six] mortality involving AMR and antimicrobial-susceptible infections.

**Figure 3.**
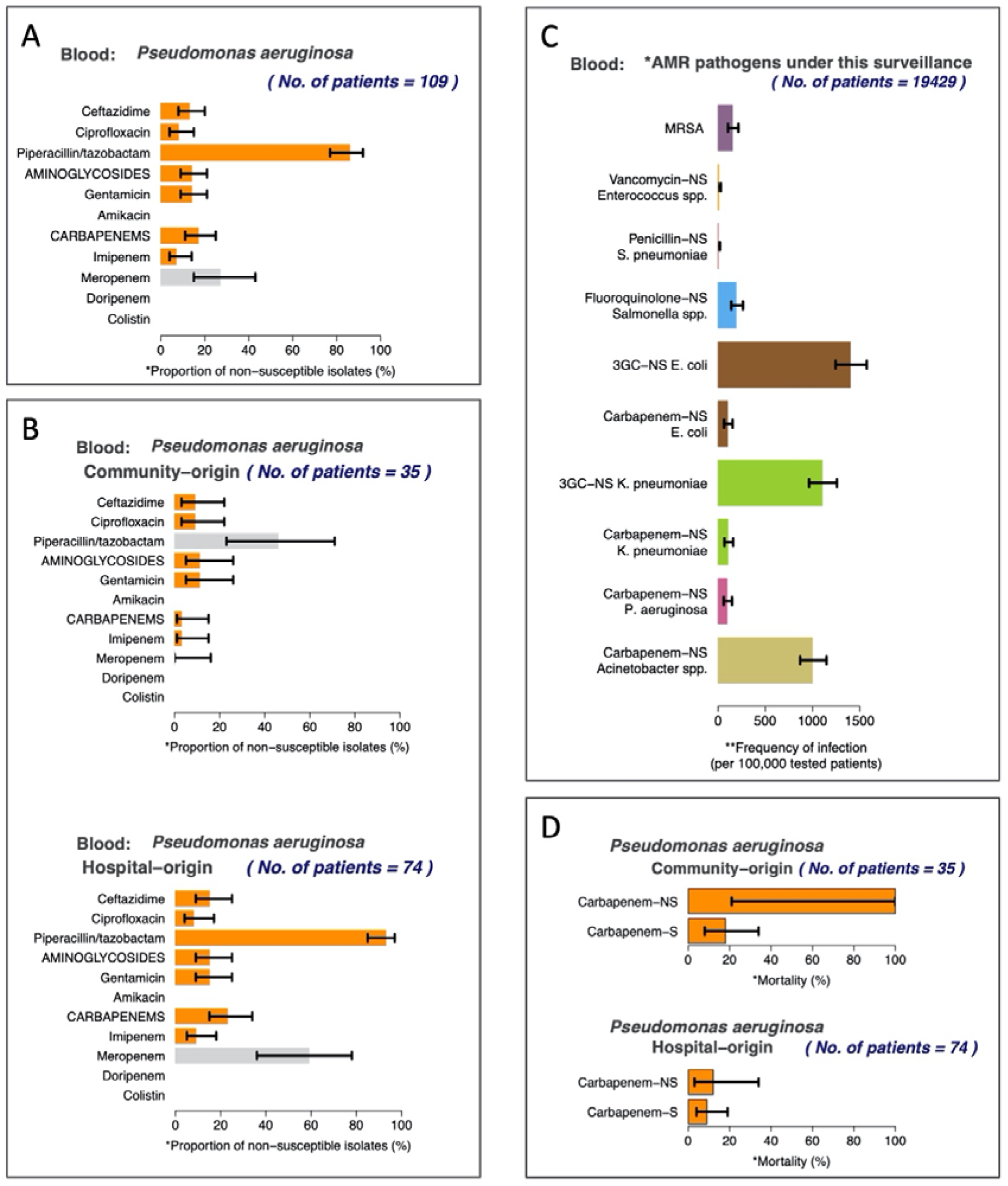
Examples of figures automatically generated by AMASS. Footnote of Figure 3. All figures are from the report (Supplementary file 4) automatically generated by the AMASS application using an example data set provided in the download package. Figure 3A represents the overall proportion of non-susceptible (intermediate and resistant) isolates in an isolate-based report (section two in the report). Figure 3B represents proportion of non-susceptible isolates stratified by origin of infection (section three in the report). Figure 3C represents the frequency of bloodstream infections per 100,000 tested patients (section four in the report). Figure 3D represents mortality involving antimicrobial-resistant and antimicrobial-susceptible bloodstream infections (section six in the report).

AMASS uses a tier-based approach. In cases when only the microbiology data file with the results of culture positive samples is available, only section one and two are automatically generated for users (as shown by the supplementary file 3, generated from the first example data set). Section three is generated only when data on admission dates are available. This is because these data are required for the stratification by origin of infection. Section four is generated only when data of culture negative specimens (no microbial growth) are available in the microbiology data file. This is because these data are required for the sample-based approach. Section five is the section four stratified by origin of infection, and is generated if admission date data are also available. Section six would be generated only when mortality data are available (as shown by the supplementary file 4, generated from the second example data set).

AMASS also generates two log files. The first log file (generated in PDF format) is for the users to validate the input data used by AMASS to generate the AMR surveillance report. It contains information such as the total number of records analysed, age distribution, number of missing values and total number of isolates per organism in the raw microbiology data file. The second log file (generated in plain text format) could be used for consultation with R users, statisticians, or the AMASS development team in case that any technical issue occurrs when running AMASS.

### Testing AMASS in seven hospitals

AMASS was tested in seven hospital in seven countries (Figure 4). The hospitals varied in data availabilities, data structure, naming of the variables, and definitions for data values (Supplementary Table 2). Overall, proportions of patients having *Escherichia coli* bacteraemia caused by 3^rd^ generation cephalosporin resistant isolates (3GCREC) ranged from 19%-85%. The incidence rates of 3GCREC bacteraemia ranged from 283 to 2737 per 100,000 tested patients among participating hospitals with available data on negative culture.

**Figure 4.**
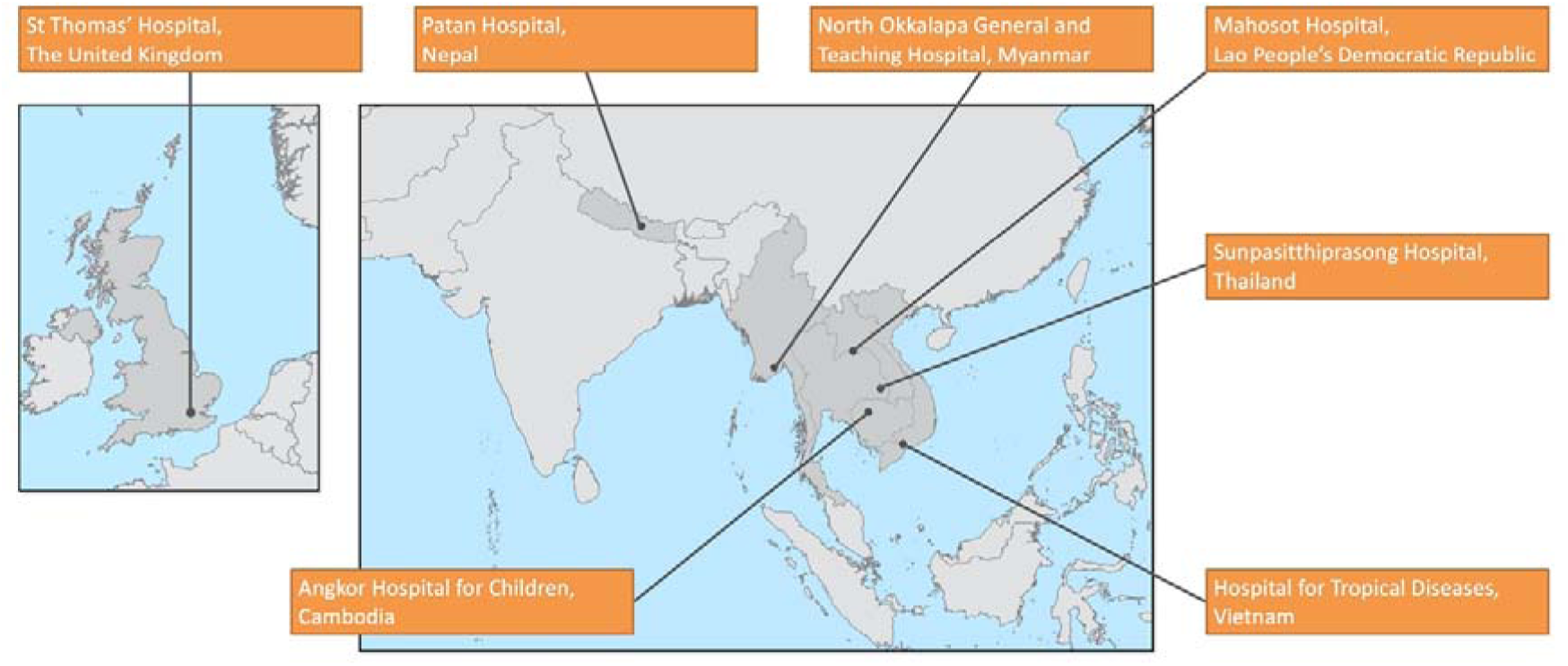
A map of participating hospitals and examples of summary data from the automatically generated AMR surveillance reports. Footnote of Figure 4. The reports and summary data from St Thomas’ Hospital, Patan Hospital, North Okkalapa General and Teaching Hospital, Mahosot Hospital, Sunpasitthiprasong Hospital, Hospital for Tropical Diseases, and Angkor Hospital for Children are open-access [28–34].

All participating hospitals deposited their output files and their data dictionary files at figshare [28–34]. Angkor Hospital for Children, North Okkalapa General and Teaching Hospital, and Sunpasithiprasong Hospital had all six sections of the AMR surveillance reports, as microbiology and hospital admission data, data on negative cultures, and in-hospital discharge outcome data were available. Mahosot Hospital and Patan Hospital had section one and two in the AMR surveillance report, as only microbiology data on positive cultures was available to test in AMASS. St Thomas’ Hospital had section one, two, three, and six in the AMR surveillance report because negative culture data were not available to test in AMASS. Hospital for Tropical Diseases had section one, two and four available, as hospital admission data was not available to test in AMASS.

Data storage systems varied across the hospitals. Angkor Hospital for Children used ACORN LIS which was based on Microsoft Access program [35]. Mahosot Hospital used a local LIS which was also based on Microsoft Access program [36]. Sunpasitthiprasong Hospital used MLAB programme [37]. St Thomas’ used ICNET^®^ (https://www.icnetsoftware.com), which is a commercial clinical surveillance software package. North Okkalapa General and Teaching Hospital used WHONET 2019 (modernized version of WHONET 5.6) [19]. Patan Hospital and Hospital for Tropical Diseases used an in-house LIS which was based on MySQL system. Inputting data dictionary files for the first time took about 1 to 3 hours. However, the data dictionary files could be stored and re-used when the raw microbiology data file or hospital admission data file was revised or updated. North Okkalapa General and Teaching Hospital used the data dictionary file from the example data set (in Example_Dataset_1_WHONET folder), which was generated to comply with WHONET exported data (in .XLSX format; Supplementary Figure 2 illustrated how microbiology data was exported from WHONET 5.6). This saved time and efforts needed to complete the data dictionaries for the hospital.

We found that the AMASS took about 1 to 3 minutes to run and automatically generate an AMR surveillance report using the local data and local hospital computers. AMASS works in Microsoft Windows 10, and with data containing a non-English (Latin and non-Latin characters) language. For example, “blood_specimen” was recorded as “Cấy máu định danh bằng máy tự động” in the raw microbiology excel file at Hospital for Tropical Diseases, Vietnam. Another example is “Male” was recorded as “ชาย” and “Female” as recorded as 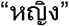 in the raw hospital admission excel file at Sunpasitthiprasong Hospital, Thailand. By completing data dictionary files, users could use AMASS to analyse the data and generate the AMR surveillance report even though the raw data were recorded in a local language.

All participating hospitals had previous experience in data verification and in AMR surveillance report validation. All participating hospitals were familiar with the recommendations of Clinical and Laboratory Standard Institute (CLSI) and European Committee on Antimicrobial Susceptibility Testing (EUCAST) on AST data verification. Additionally, all participating hospitals were familiar with validating summary data by comparing the summary data automatically generated by the AMASS application with manual calculations.

## DISCUSSION

We developed an open-access, offline and easy-to-use application, AMASS, which allows hospitals, especially in LMICs, to automatically generate AMR surveillance reports from routinely collected electronic microbiology data. Seven hospitals used AMASS, and shared their reports and summary data files by depositing them in an open-access data repository website. We propose that routine generation and sharing of AMR surveillance reports (i.e. cumulative antimicrobial susceptibility reports or antibiograms) in open-access data repositories from hospitals with a microbiology laboratory should be supported worldwide [38].

AMASS empowers data sharing by reducing the time and effort needed to prepare the summary data and reports without increasing the risks of breaching individual patient data confidentiality. This is because AMASS can analyse the data and generate the reports promptly without needing to transfer raw data files to any party outside of their hospital. The auto-generated AMR surveillance reports are in PDF format which can be reviewed and shared locally and immediately. This is important to improve understanding of the local AMR burden and to inform local patient management. The PDF report and summary Excel files contain no individual patient-level information and can be shared with national and international organizations to support action plans on AMR. Any attempt to compare AMR surveillance reports among different hospitals should be done cautiously, as factors including the type of hospital, blood culture utilization rates and practices, and patient characteristics may influence the estimated prevalence and incidence rates of AMR infections [12–14]. An AMR surveillance report should clearly present denominators used to calculate incidence rates. Selecting an appropriate denominator that represents the local setting is important for estimating incidence rate of AMR, but remains challenging [39]. AMASS calculates the total number of tested populations and used as the denominators in Section four and Section Five of the output report based on the WHO GLASS recommendation, and this denominator will not be available without data on negative culture.

AMASS uses currently recommended analytical approaches to generate the summary data and reports. All of the code is open-access, verifiable and modifiable. AMASS can also be expanded and tailored based on the local requirements. For instance, notifiable bacterial infections that are important for a local setting can be included by revising the R code provided in the AMASS package. Analytical methods can also be constantly updated and improved over time.

AMASS is an add-on automatized report generating that can be easily used, even by hospital users with limited information technology kills. It does not replace WHONET, LIS, quality assurance programmes, or antimicrobial surveillance systems (including the WHO GLASS). AMASS therefore differs from ‘Macros and Excel Reports’ functions of the WHONET, which allow users to regularly generate reports on screen and export data as Excel files using macro language (i.e. a series of written study parameters generated automatically through the WHONET user interface grouped together) [19]. In contrast AMASS is designed to automatically compile summary results of multiple organisms into a single report. The current version of AMASS does not support quality improvement or alert individuals to unexpected antibiogram results, which WHONET is capable of [19]. Thus, while WHONET is an appropriate choice for the main software package for microbiology laboratories in LMICs to capture and store microbiology data, and to export data for the WHO GLASS [19], AMASS provides significant additional functionality [19].

Verifying individual AST results and validating summary data in the reports (including those generated by AMASS) remain major barriers to the generation of accurate surveillance data on AMR. Only verified AST results on each patient’s isolate should be used in the data analysis [12–14]. WHONET and many LIS include functions that automatically check the results to ensure that they appear reasonable or alert the users to confirm unusual results (e.g. amikacin resistance coexisting with gentamicin and tobramycin susceptibility in *E. coli* is unusual and data on such cases should be verified) [12–14,19]. In addition, it is important to validate the calculations of any analytical software used to generate the summary data and reports. We recommended users of AMASS perform manual validation (such as printing a line listing of all isolates of the species to cross-check with the reports), particularly when the program is used for the first time [12–14]. Users should also understand that the precision of the estimate is based on the sample size, which is illustrated by estimated confidence intervals. In addition, representativeness of the summary cAST data could be impacted by the sampling strategy used, particularly if blood culture is mainly collected from patients with treatment failure or prolonged hospitalizations [11,12,14]. Nonetheless, generating AMR surveillance reports from existing microbiology database could be a first step for hospitals in LMICs to understand and validate their own data, and support quality control programmes at their local level.

Local hospitals must not be discredited because of the statistics in any AMR surveillance report, as possible criticism could be a barrier to data sharing [40]. Negative criticism on the data shared by the local hospitals needs to be avoided. It is inevitable for errors to occur in any data set especially in settings with limited resources and experience on quality assurance and data validation. Tools for data validation should be provided for local hospitals, and the statistics in an AMR report should act as a guidance on the direction of improving data quality control, data validation, and infection prevention and control. Credits should be given to the hospitals that share their local AMR data and reports at an open-access data repository platform. The data repository platform, such as figshare, allows flexibility on updating the data and revising the AMR surveillance reports with appropriate version controls by the local hospitals. Publishing open-access AMR data on these platforms provides a digital object identifier (DOI) that should be used for data citation [41].

### Potential issues and limitations

AMASS has a number of limitations. Firstly, AMASS is not applicable for hospitals that only store data on paper forms. Secondly, AMASS cannot work with raw microbiology data file that are not in wide format or combine many files in multiple formats; for example, the raw microbiology data file where each row contains data of each antibiotic susceptibility result for one single specimen. Thirdly, the current version of AMASS can only analyse microbiology data that includes antimicrobial susceptibility test interpretive categories (susceptible, intermediate, resistant) based on guidelines that the local hospital uses. Fourthly, AMASS cannot automatically validate the reliability of data that are imported into the application and used for analysis. Data verification and quality checks will be included in the future versions of the application. Fifthly, AMASS was tested with only few data sets that included non-English languages, and further testing on data with other languages may be needed. The current version of AMASS has been tested in Windows 10 and 7 and may not work under different operating systems. Finally, the current version of AMASS cannot provide options to export the AMR surveillance report in different formats (i.e. in document or text formats). However, users can re-use the summary statistics, which are saved in CSV format under the ResultData folder.

## CONCLUSION

AMASS can be used to support hospitals with microbiology laboratories in analysing routinely collected data and generating reports with minimal resource and expertise required. This may empower individual hospitals to contribute to the understanding and actions on AMR at local settings, and maximize the utility of the local data.

## Data Availability

The data that support this study are open-access in figshare at:
https://doi.org/10.6084/m9.figshare.12000225.v1
https://doi.org/10.6084/m9.figshare.12000222.v1
https://doi.org/10.6084/m9.figshare.12000237.v1
https://doi.org/10.6084/m9.figshare.12000231.v1
https://doi.org/10.6084/m9.figshare.12000240.v1
https://doi.org/10.6084/m9.figshare.12000249.v1
https://doi.org/10.6084/m9.figshare.12000252.v1

## Acknowledgements

We thank all of the hospitals participated in the AMASS project. We thank Dr. Thet Wai Nwe from the Central Epidemiology unit of Myanmar; and Dr. Htay Htay Tin from the National Health Laboratory of Myanmar for their comments on AMASS. We also thank Dr. Kyi Soe, who is the Senior Medical Superintendent of North Okkalapa General and Teaching Hospital for his support for the AMASS and hosting the AMASS training workshop at the hospital. We thank Professor Frank Smithuis and Dr Aung Pyae Phyo from MOCRU for their supports on the AMASS project. We thank Professor Paul Newton and Professor David Dance from the LOMWRU for their supports on the AMASS project. We thank Ronas Shakya from Patan Hospital, Nepal for technical assistance. We thank Joe Hessell from Diagnostic Microbiology Development Program for comments on AMASS. We thank Chotipong Chaisiripreeyakul, Techat Totemchokchaikarn, Sitthinun Pasirichusaree, and Papitchaya Achavakulthep for the tutorial videos for AMASS.

## Author contributions

C.Lim and D.L. conceptualized, designed, and developed AMASS, and wrote the original draft. T.M., V.C., M.T.A., A.K., P.Te., R.B., and L.N.P.H tested AMASS, curated data, and reviewed reports generated from AMASS using local data. J.S., P.Tu., E.A., R.v.D., H.N.L, C.Ling, S.H., S.I., S.D., T.W., V.H., W.S., Y.L.M., T.L.V., H.H.H., M.M., M.V., B.B., J.E., S.J.P., G.T., N.P.J.D., and B.S.C. reviewed and interpreted the reports. All authors participated in the drafting, editing and revising the final draft.

